# Understanding the cultural environment of the outpatient care setting for patients with dementia receiving cancer treatment: a qualitative study

**DOI:** 10.1101/2022.04.20.22274020

**Authors:** Naomi Farrington, Katherine Dantanus, Alison Richardson, Jackie Bridges

## Abstract

**Introduction:** People with dementia have poorer cancer outcomes than those without, and experience inequalities in access to, and quality of, care. Outpatient environments, where radiotherapy, chemotherapy and immunotherapy cancer treatments typically take place, have largely been excluded from research. This study was conducted to understand provision of treatment and support and experiences of care for people with dementia undergoing cancer treatment in the outpatient setting.

**Materials and methods:** Using observation, interviews and document analysis, data were collected to scrutinise the cultural environment of ambulatory care, comprising the physical fabric of the care setting; interactions, behaviours and perceptions of those in the care setting; and the organizational, clinical and interactional processes involved in care delivery. The study was conducted in the outpatient oncology departments of two large teaching hospitals in England between January 2019-July 2021.

**Results:** Data were gathered from a wide range of sources, including 15 hours of observation, and interviews with patients (n=2), caregivers (n=7) and staff (n=20). Evidence from this study suggests the cultural environment of the outpatient care setting reflects and supports the standardised processing of people for cancer treatment. Dementia introduces a wider set of care requirements not catered for by this standardised treatment model and associated processes. Data showed the needs of patients with dementia could be addressed most effectively when individualised, as opposed to standardised care, was offered.

**Conclusion:** There is work to be done in outpatient cancer services to ensure responsiveness to individual patient need. This could be achieved by having an established way (or ways) of eliciting needs, preferences and expectations, a belief that a person’s needs and expectations are legitimate, and that effort should be made to address them, with the ability to accommodate these needs and expectations.

**Patient or public contribution:** patients and caregivers were involved in the study design and development of study materials including the interview topic guide. They also assisted with discussion and clarification of study findings.

## Introduction

People with dementia have poorer cancer outcomes than those without [1, 2]. A scoping review [3] identified limited interventions to support older people with complex needs having cancer treatment. Good quality evidence is lacking regarding the implications of comorbid cancer-dementia for people receiving cancer treatment, and little information exists about how the needs of this population are managed by healthcare teams [4].

People with dementia experience inequalities in access to, and quality of, care [5, 6]. There is a focus on the disease instead of the whole person, and a focus on a single disease alone, which can be disadvantageous for people with multimorbidity. People with dementia frequently feel denied, ignored, or experience discrimination in healthcare [7] either as a direct result of the stigma associated with the diagnosis, or through indirect mechanisms such as failure to provide inclusive services. It is increasingly recognised that a tailored approach to multimorbidity is required [8] enabling individual preferences and circumstances to be addressed.

Efforts have been made to ensure people with dementia have access to services that meet their needs by establishing care standards [9]. A body of work exists on improving care environments for people with dementia in hospital as inpatients or in residential care [10,11]. Outpatient environments, where radiotherapy, chemotherapy and immunotherapy cancer treatments typically take place, have largely been excluded from these efforts, although evidence is emerging [12]. Care and treatment in an outpatient environment are different from that of an inpatient stay. Outpatient care involves a series of discrete interactions; patients attend the service for specific appointments, such as consultations and treatment, and in between return home. In addition, the patient (or caregiver) is expected to accept responsibility for coordinating appointments and treatment, as well as monitoring their own health and wellbeing for treatment-related toxicities. Outpatient treatment, while reducing time at the hospital, brings other challenges.

This study investigated the provision of treatment and support and the experiences of care for people with dementia undergoing cancer treatment in the outpatient setting. It aimed to establish an empirically-based conceptual foundation to inform development of innovations in service delivery, and improve the way treatment and support is offered to this group.

## Materials and Methods

Data were collected to scrutinise the cultural environment of outpatient cancer care characterised through the interactions, behaviours and perceptions of those in the care setting; organizational, clinical and interactional processes involved in care delivery; and physical fabric of the care setting. Data were used to identify principles and characteristics that constitute ‘good care’, understand barriers and facilitators, and identify aspects amenable to modification to meet the needs of this complex population. Study design was influenced by focused ethnography, which enables concentration on a distinct issue or shared experience in a specific setting [13,14,15]. It has been used successfully in nursing research [16,17], particularly when the researcher is known and trusted, and holds a ‘privileged observer’ position [18]. Two of the researchers (X and X) were nurses in departments involved in the study. Insights afforded to them by their clinical roles were invaluable in understanding care delivery.

The study was conducted in the outpatient oncology departments of two teaching hospitals in England. Data were collected January 2019-July 2021. Observation, semi-structured interviews and examination of patient case notes were used in a focused manner by two researchers (X, X). Participants included: patients, informal caregivers, and healthcare staff (see **Table 1**). The process is described below; however, the study comprised more than the recruited participants and interviews. Ethnographic data covers a broad ontological range from ‘hard, objective’ documents, to ‘soft, subjective’ memories and experiences [19]. The field researchers (X and X) internalised the research aims for the period of data collection, and interpretation and understanding continued outside assigned research time. The findings reflect a wider data field than the formal data collection opportunities described.

**Table 1.** Inclusion and exclusion criteria for interviews.

|  | Inclusion | Exclusion |
| --- | --- | --- |
| **Patient participant** | ○ Adult, aged over 18;<br>○ Diagnosis of any cancer;<br>○ Undergoing cancer treatment (radiotherapy, or chemotherapy or other SACT delivered via any route) OR have finished treatment within the last 6 months;<br>○ Cancer treatment administered in outpatient care setting;<br>○ Diagnosis of dementia of any type;<br>○ Mental capacity to decide to take part in the study. | ○ Acute or critical illness;<br>○ Inability to communicate choices and preferences either verbally or non-verbally;<br>○ No confirmed diagnosis of dementia;<br>○ Cognitive impairment as a result of aetiology not related to dementia. |
| **Other participants** | ○ Adult, aged over 18;<br>○ Informal carer of patient participant OR healthcare professional/ NHS staff involved in care & management of patient participant or other patients with dementia having cancer treatment. |  |

### Patient participants

Patient participants were purposively sampled to participate in interviews and observation. The study invited participation from people with a dementia diagnosis who were receiving radiotherapy, or systemic anti-cancer therapy (SACT), or who had completed treatment within six months. Patient participants were identified through their clinical teams, and agreement gained for researchers to approach them (face-to-face where possible, via telephone during the COVID-19 pandemic). Participation in each of the three data collection methods was not mutually exclusive, nor was it mandatory; patients could be interviewed, observed, and consent to document analysis, alternatively they could only be observed, or only interviewed. Document analysis was undertaken if a patient participant consented to this at interview.

### Caregiver participants

Caregiver participants were family or friends involved in supporting a patient through radiotherapy or SACT. They could participate alongside the patient they were supporting, or alone. As with patient participants, they were identified by the clinical teams, and gave agreement for the researchers to contact them.

### Healthcare staff participants

Healthcare staff participants included: oncologists, nurses, allied health professionals, support workers, and management and administrative staff. Purposive sampling was used to recruit staff involved in the delivery of treatment and support to patients undergoing radiotherapy or SACT. They did not have to be directly providing care to a patient participant.

### Data sources

Interviews took place in a private area of the department, or a participant’s home (an option offered to patients and caregivers). A topic guide, developed with Public and Patient Involvement (PPI) volunteers, provided discussion prompts. Participants were invited to describe their experiences. Subsequent interview questions covered the treatment environment, factors people found challenging, and what they found helpful. Interviews were digitally recorded and transcribed verbatim. Where patient participants gave permission, data were extracted from case notes about their journey within and beyond cancer care, and the organizational, clinical and interactional processes involved in care delivery.

During general observations, attention was paid to the environment, behaviour and staff-patient interactions, with a focus on delivery and experiences of care. Focused observations allowed a detailed study of discrete experiences, such as pre-treatment consultations. Attention was paid to the environment in which care was experienced and delivered, the behaviours of the people involved, and the organisational processes enacted. Descriptive and reflective field notes were captured on an observation record form.

### Ethics

Ethical approval was gained from South Central–Berkshire Research Ethics Committee (18/SC/0590). Informed consent was secured for interviews, case note access and focused observations. Verbal explanation of the researchers’ presence was provided when requested during general observations. The UK Mental Capacity Act [20] was used to guide researcher assessment of people’s capacity to decide to take part in the research. COREQ reporting criteria checklist for interviews and focus groups [21] informed the writing of this paper.

Analysis was conducted concurrently with data collection [22,23] by two researchers (X, X) and discussed and iterated with the wider team once the full data-set was available (X, X, X). Based on the constant comparative methodology of grounded theory [24,25] the process was as follows:

1. Initial coding (categorising data)
2. Focused coding (concentrating on significant/frequent codes)
3. Theoretical coding (developing relationships between codes)
4. Memo-writing (analysing ideas about codes)
5. Theoretical saturation
6. Sorting and integrating memos

Analysis followed the same pattern regardless of data source type and involved looking for patterns and relationships, as well as inconsistencies and contradictions. Several themes were identified, refined and reviewed to include those that captured the story being told by the data [26]. Data collection ceased when the researchers had achieved adequate depth of understanding to build theory [27].

## Results

Data were gathered from observation (15 hours), interviews with patients (n=2), caregivers (n=7) and staff (n=20) (see **Table 2**), document analysis and informal discussions. Interviews lasted between 10 and 42 minutes. Four patients approached declined participation.

**Table 2.**
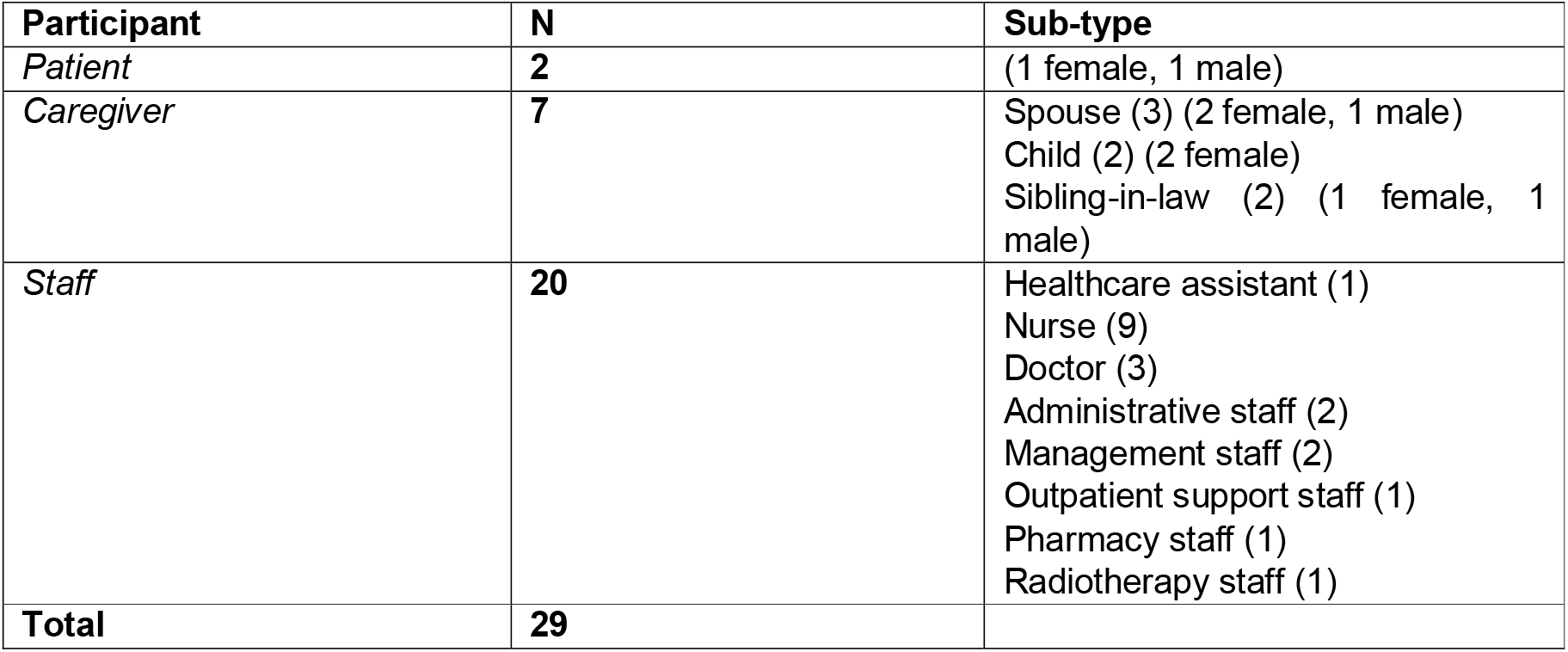
Participant details.

To set the scene for patient/caregiver experiences and the cultural setting that shapes them, we begin by outlining how relevant services were organised in the departments providing treatment. We then elaborate on the key steps that occur during a patient journey: ‘*attending the hospital’*; ‘*the consultation*’; ‘*treatment*’; and ‘*at home*’, illustrating each stage. Pseudonyms are used to protect anonymity.

The SACT department included outpatient clinics where patients attend for consultations, clinical areas where patients attend for blood tests, and treatment areas. Prior to the COVID-19 pandemic, patients tended to see a clinician face-to-face for a consultation (doctor, nurse practitioner or pharmacist), and have a blood test, before either collecting oral treatment from the hospital pharmacy, or returning on a subsequent day for intravenous treatment. This system changed during the pandemic to reduce hospital attendance; most consultations were conducted remotely via telephone or video, blood tests done at home or at GP surgeries, and some treatments administered at home. However, the overall process remained unaltered during and beyond 2020, regardless of setting: patients generally need a consultation, a blood test, and a treatment appointment. Treatment in the radiotherapy department was not altered by the pandemic, as radiotherapy can only be provided on a hospital site. Patients attended for a pre-treatment planning appointment, with a CT scan to determine the specificities of treatment. Patients were then given a treatment schedule, either for a one-off treatment or for a course of treatments. Radiotherapy and SACT treatments were sometimes given concomitantly. In both areas, patients were discharged home after treatment, with contact details for a 24-hour acute oncology service. Interactions between patients and the oncology service were organised to enable diverse patients to progress along these standardised pathways.

### Attending the hospital

Participants cited difficulties with attending for hospital appointments. Services were organised and delivered in a standardised way which could create problems for patients and their accompanying family members. The sensory experience of being in the clinic was shaped by the built environment and the busyness of services, and patients found being there unpleasant and disorienting. These negative experiences were exacerbated by frequently experienced long waits. Alternatives to visiting the hospital were welcomed by some when they were made available, but did not suit everyone’s needs.

Participants found it difficult to arrive on time for set clinic appointment times, especially in the morning:

> “*Getting up, breakfasted and all ready and then getting up here and finding a parking space, it’s a bit of a panic if you’ve got to be here for 9*.*20am*.” (Annabel, wife of person with cancer and dementia)

Long waiting times also created problems if people could not be seen in a timely way once they had arrived or had to wait between different parts of the appointment:

> “*He gets confused by the waiting. Well, he has, I’m not quite sure whether he’s just not used, he’s not used to NHS and being ill and everything like that because he’s not an ill person previously, so he can’t understand why he has an appointment for say, I don’t know, 9 o’clock, and he’s still sitting there, you know, sometimes, he could be sitting there an hour later*…” (Emily, wife of person with cancer and dementia).

The noisy and busy nature of the clinic environment aggravated the experience of long waits and features of the built environment heightened patients’ disorientation:

> “*It’s very busy, it’s very noisy, it’s very complicated, the doors look the same. Patients can get lost in sitting in a room with doors looking exactly the same everywhere*.” (Tracy, nurse specialist)

> “*Noisy: staff calling out patient names for height and weight, appointments, treatment, numbers for bloods, phones ringing, bin lids slamming. Very difficult to tell from which direction the noises are coming from, so sometimes patients are unaware where to go when they are called*. (General observation of SACT clinic area)

Staff told us that sometimes the long waiting times were an inevitable part of attending the hospital for cancer treatment:

> “*… when patients come to the hospital there are a number of waits and that could be waiting for blood results, waiting for going in for treatment, seeing the doctor, whatever those waits are, but there are waits built into the service…. But actually, I don’t think until you actually come to the hospital and you are actually involved in those waits you can really actually realise how long some of those waits can be*.” (Maisie, SACT management team).

Efficiently accommodating large volumes of patients was managed by “overbooking” clinic slots, that is allocating more than one patient to be seen at a time. This meant the capacity of clinic staff was exceeded and patients were inevitably delayed beyond the time of their booked appointment. Delays also occurred if patients earlier in the schedule took longer than the scheduled time for their appointment, a frequent occurrence as actual patient need was hard to predict and accommodate in the booking system where all clinic slots were the same length.

Patients and caregivers told us they appreciated having fewer hospital visits, and we found examples where a reduction in visits had been made possible by appointments held over the telephone:

> *“So we come here on Thursday, and the treatment is the following Tuesday. But now, the doctor’s just said [my husband will visit the hospital] every 6 weeks, because he’ll get a phone call on the third [week instead of a hospital visit], so that makes it easier*.*”* (Emily, wife of person with cancer and dementia).

But what presented as a welcome solution for some, did not fit with what others needed, highlighting how service provision needed to be individualised. Imogen, for instance, found that her father, who had advanced dementia and no longer spoke, was repeatedly sent letters inviting him to a telephone appointment, requiring her to contact the department numerous times to explain that her father was unable to use the telephone.

The frustrations and disempowerment reported by patients and caregivers were the result of a mismatch between the standardised way that services were organised and the needs and preferences of patients and caregivers. Dementia added another layer of complexity: it was harder to get to appointments on time, long waits were more onerous, and the physical environment of the clinic was more disorientating. Dementia also complicated uptake of alternatives such as telephone appointments. Our findings also cast light on the impetus behind the way that services are currently organised and the primacy of organisational efficiency, at the expense of patient experience.

### The consultation

Experiences of consultations during clinical visits tended to be shaped by the degree of continuity between one visit and the next, and by the extent to which the consultation aligned with patient concerns that may extend beyond cancer. Specialist roles in nursing and in older people’s care were identified as having the potential to improve the quality of consultation in relation to actual patient need and to improve continuity between appointments.

Interviewees reported that continuity was disrupted when the staff in the consultation changed between visits:

> “*Yeah, I think just, yeah, we saw different doctors every time, and I know that was quite hard. But it was a case of who’s doing what? OK, who’s chasing the scan? Is that going to get done? It was just that really, and tying it all together*.” (Melanie, daughter of person with cancer and dementia).

Case note analysis confirmed high numbers of staff to interact with as a potential challenge. For instance, Katherine had 47 discrete interactions with 24 different clinicians in oncology over 13 months. As the quote above illustrates, caregivers were not always assured that there was one professional with the overview of the case who was making sure that all the elements of care and treatment were appropriate, integrated and consistent. The impact of dementia on communication quality and on memory, in addition to high turnover of clinicians, placed additional burdens on family caregivers who felt responsible for ensuring that information was transmitted and received during consultations and retained afterwards. Melanie found the lack of consistency with a central person difficult, as she found herself stuck in the middle during consultations with healthcare professionals who had not met her father before:

> *“I think the initial meeting…It didn’t seem, it wasn’t rushed, but I had to audio record [the meeting], because things got lost, and Mum would hear what she wanted to hear, Dad wasn’t listening at all, I was trying to interpret for the doctor and understand Dad, because he was so bad at the beginning, and then we had the nurse in there as well, saying things, that was a bit too much, that was overwhelming*.”

Emily, supporting her husband with cancer and dementia, found it difficult to remember the names of the different doctors they had seen, referring to one as “*Dr, whose name begins with [x]*”. Her husband found the lack of continuity problematic:

> *“He’ll say afterwards, or later on, ‘I keep seeing different people’, and he finds that a bit confusing*.*”*

The high numbers of patients needing to be seen meant that individual consultants could not see all their patients every time they came to clinic. Patients who were viewed as continuing well on treatment and having no additional need would instead be seen by others, such as advanced nurse practitioners. The pressure on clinics and their clear remit for cancer also constrained the topic of discussions to the cancer, with more pressing concerns of patients and caregivers not being aired. During a conversation with Paul, who had cancer and dementia, and his wife Annabel, it became apparent that the couple’s primary concerns were not about cancer:

> *“Wife becomes very tearful and says that it has been a difficult month. Says that patient’s sister has died at the beginning of March from vascular dementia, and that the patient has been diagnosed with vascular dementia yesterday. In addition, he had a driving assessment and is no longer safe to drive. On being reminded of all this the patient starts to become visibly tearful. Wife goes on to describe that the diagnosis of vascular dementia arrived by letter yesterday with no notification or suggestion of follow up. She appears very upset by this*.” (Field note).

Paul’s diagnosis of vascular dementia, and the fact that he could no longer drive, were having a greater impact on Paul’s life than his well-controlled cancer. The consulting clinician was sympathetic, but the primary concern was assessing treatment toxicities and prescribing further treatment, as this was where the clinician’s expertise and responsibilities lay. Staff participants acknowledged that cancer would not be the only health issue for many patients, and suggested that involvement from specialists in medicine for older people could be beneficial, and allow conditions such as dementia to be addressed alongside cancer treatment:

> *“So you also have a geriatrician involved in their care so you are looking more holistically at everything that is going on with them because they are a group of older people so they don’t just have, 90% of the time they don’t just have a cancer they have other medical problems going on”* (Tracy, specialist nurse)

Caregivers consistently cited the specialist nurse role as key in promoting continuity of cancer treatment:

> “*Yes we’re all sorted now, I know that I can ring [specialist nurse] to find out anything about my dad’s blood or blood test or anything like that at all I know any questions or anything I can ring and if [specialist nurse] doesn’t know she’ll find out and then she’ll ring me back. So since I found that I have got someone that I can speak to it’s a lot better now*.*”* (Imogen, daughter of patient with cancer and dementia).

Specialist nurses were able to operate outside of the standardised service processes to address the complexities that dementia added. For instance, Sheila (sister-in-law of person with cancer and dementia) appreciated the fact that a nurse rang her after consultations to summarise the content. Melanie welcomed the hospital arranging for appointment letters to be sent out both to her and her father’s care home, so she was kept up-to-date with her father’s treatment.

These findings highlight that high volumes of patients in relation to consultant capacity were managed by substituting consultants with other staff members. This strategy disrupted continuity between visits, creating confusion and stress for patients and caregivers, and impacting negatively on relationship quality and information transmission. Nurse specialists had the autonomy to operate outside of the standardised service to promote continuity. In addition, the high pressure on clinics and the service focus on cancer alone constrained opportunities for patients and caregivers to raise non-cancer issues, however pressing they were for health and well-being. The broader focus of medicine for older people specialists was cited as having the potential to address such issues, but was not available to patients in the study setting.

### Treatment

Patient experiences of receiving treatment were shaped by the readiness of the department to accommodate their dementia at the same time as cancer treatment. We found variation in whether or not departments were notified in advance that someone had dementia but also in the extent to which effort had been put into making the service “dementia-friendly”.

At one study site, the booking team highlighted people with dementia to radiotherapy staff so that the appointment could be planned accordingly. Staff in other departments told us they were not always aware that patients had dementia before they arrived:

> *“…because the problem is quite often we’re just chasing our tails. We don’t know any issues until we’ve already booked the patient and they walk in the door and suddenly we find that it’s not suitable anymore*.” (Louise, administrative team)

In the radiotherapy department, radiographers with specialist dementia training oversaw the care of patients with dementia. Before the appointment they contacted family or friends to see how best to support the patient. They then met the patient at the planning appointment, giving them a chance to identify what adaptations might be needed to the treatment plan. In departments without dementia champions or where links were not in place with the hospital’s dementia nurse specialists, staff were required to work in a more reactive way but struggled to provide the quality of care they felt was needed. If the patient needed more time with staff because of their dementia, staffing levels did not allow for this:

> “…*when you are the nurse who is trying to concentrate and with chemo you’ve got to really concentrate on checking the bags, the dosage, and sometimes when they’re called away for other things there’s a risk of error and this is what the nurses say, and you’d hear them say it and I know awful, oh God they said Dorothy is in today. Because she was so labour intensive and it’s not the physical side it’s the emotional side because she will be constantly saying, why am I here, why am I here. Well again if you are working and you are trying to concentrate and she’s constantly, they don’t allow for that side of it for nursing staff*.’ (Natalie, healthcare support worker)

Some staff members were not confident they had the skills for supporting people with dementia:

> “*One of the little nurses in there even turned around and said I’ve never spoken or dealt with a person with dementia before so I don’t really know what I’m supposed to do*.” (Imogen, daughter of person with cancer and dementia)

> “*I probably wouldn’t be confident in knowing where to go and how to find out specifically related to the dementia side rather than the oncology side*” (Tracy, specialist nurse).

Accommodating someone’s dementia while delivering their cancer treatment depended on the motivation, knowledge and skills of staff in individual departments. Where departments were ready for patients with dementia, an individualised approach could be planned. Where departments were not ready, the resulting standardised approach to staffing and care meant that patient experiences and potentially outcomes were negatively affected.

### At home

Our findings highlight the key role family caregivers play in supporting people with dementia through their cancer treatment, especially in the home setting, and in turn, the ways in which family caregivers can be supported in their role by cancer services. Doctors told us that the presence or absence of supportive family at home affected their decision to give certain treatments:

> “*I think it’s a safety thing from our point of view if we know that we’ve got a family that’s going to remind that patient what the diagnosis is, what the plan is, what the treatment is, then we would feel much safer perhaps prescribing a more intensive treatment that suits them physically than for a patient that has nobody at home that’s not going to remind them to take their tablets…*.*to come in when they’re unwell*.” (Teresa, doctor)

Caregivers found home care challenging during cancer treatment, especially managing medicines, a role made necessary by the memory problems that accompanied dementia:

> “*I have to do all his pills because otherwise they might get forgotten. He has five pills in the morning, two pills at 11am, another pill after 12pm and then his evening pills he’s got three more during the evening, one with his meal and two before he goes to bed. So it’s pill, pill, pill all the time*.*”* (Annabel, wife of person with cancer and dementia).

Staff recognised their role in supporting caregivers, including giving them more advance information about the whole treatment plan than would usually be provided:

> “*Yes, I guess that as nurses we’ve got to care for the patient and also their support network and their loved ones so we’ve got to really make sure that we’re looking after the whole package because if a carer is struggling they’re not going to be able to be there for the patient*.” (Leila, nurse)

> “*A lot of people don’t appreciate actually 12 cycles could mean you are into February next year and it doesn’t quite compute until you see it on paper and go oh my God. So if you can give them that right from the beginning then they can work to a plan*.” (Louise, administrative team)

This advanced notice enabled families to plan ahead but whether or not this information was provided depended on the discretion of the individual staff member.

Our data showed that the needs of patients with dementia (and those of their caregivers) could be addressed most effectively when non-standardised (individualised) care was offered. Evidence from this study suggests that the cultural environment of the outpatient care setting reflects and supports the standardised processing of people for cancer treatment. Dementia introduced a wider set of requirements that were not catered for by this standardised treatment model. The discussion below considers how healthcare systems could address this gap in provision.

## Discussion

This study investigated the provision of treatment and support, and experiences of care for people with dementia having cancer treatment in an outpatient setting. Attending the hospital to arrive at a scheduled time and waiting to be seen was stressful for patients and caregivers. Not seeing the same staff member at every visit disrupted informational and relational continuity [28] and was burdensome for caregivers whose role become one of mitigating for lack of continuity. The focus of consultations on cancer limited opportunities to raise other concerns. If treatment departments were not readied to treat people with dementia, staff stress and poor patient experiences resulted. The findings illustrate how routine, standardised approaches to the organisation and delivery of cancer care and treatment were in tension with the needs of people with dementia. A personalised approach was possible when staff had skills, discretion, flexibility and resources to plan ahead, to elicit what an individual and their caregiver needed, to accommodate health needs beyond cancer and modify plans and treatment to fit individual needs. Participants indicated that nurse specialists, dementia champions and specialists in medicine for older people were helpful “anchors” in this regard, as they had broader skills and knowledge and took into account the totality of a patient’s needs. But there were clear variations in the extent to which such roles were routinely accessible to people with dementia and their caregivers.

Balancing the needs of a person with dementia and the requirements of a cancer service is a recognised challenge [29]. High quality dementia care is dependent not just on the efforts of individual workers at the point of care, but also on the extent to which the wider infrastructure enables high quality care to be delivered [30, 31]. Systems that enable this can be described as *responsive* [32]. This concept conveys how health systems can dynamically respond to changing needs and encounter the patient as a genuine partner, identifying and meeting each person’s needs and expectations in the context of their personal goals and preferences [33]. Although responsiveness can exist alongside standardisation [34], the findings from this study suggest that this is a challenge.

Standardised packages of cancer care specify a predictable patient journey through the system from diagnosis to completion of treatment [35]. A standardised approach allows for the staff and technologies that deliver treatment to be assembled in place at the right time so that care is delivered as planned and the patient moves on, while the assemblage of staff and technologies moves efficiently to the next patient [36]. Movement is timetabled on the assumption that every patient needs an appointment of equal length, but our findings show that this rational plan is disrupted as patient needs are not uniform. Although patients further down the list are ready for their appointment, they must wait until the appointment is ready for them, suggesting that the distribution of waiting time coincides with the distribution of power [37]. We observed the practice of organising clinic visits in a “hyper-rational” way; allocating more than one patient to be seen at a time, exceeding the capacity of available staff and resources, thus increasing the likelihood of waiting and inconvenience to the patient and caregiver. Any resulting poor experiences for the patient and caregiver are not accounted for when a rational model of work dominates. Primacy is given to the need to keep the clinic running efficiently: patients wait so staff don’t have to.

Standardisation may contribute towards efficiency, but is not automatically equated with quality [38]. In spite of rational approaches being used to organise care delivery, there can be high variation and unpredictability at an individual patient level [36] and our findings illuminate how dementia can be a source of unpredictability. Dementia may also constrain a patient’s ability to self-manage outside the hospital environment, which is key to outpatient cancer treatment. The dementia is inconvenient in a bureaucratic system because of its potential to disrupt patient flow through the system and interfere with efficient use of resources.

Health care work such as cancer treatment is increasingly specialised, with different parts of the care trajectory being handled by different teams of people distributed across time and space [35]. This fragmentation means patients can only present a particular element of their health in the context of each individual encounter with a service. This allows the clinic to keep to time, but the outcome for the patient is that only their cancer is a legitimate topic of interest. The structural lack of coordination described here is known to exacerbate treatment burden [39]. Our findings reveal the impact of this fragmented experience for patients and caregivers. In the absence of professional roles that provide the anchor-point, they must provide the continuity of information and care management that otherwise feels absent.

The fragmented experience and focus on cancer alone marginalises the dementia and renders it invisible in standardised approaches to cancer care. As such, decision-making in the oncology clinic can only relate to cancer, even if patients have more pressing health concerns beyond this. The dementia therefore seems less relevant, and is less likely to be part of the conversation, which prevents highly specialised practitioners from having a full understanding of the patient’s body and personal experience [35]. In turn, this inhibits how responsive they can be to patient need.

Our findings identified key system issues that impeded the capacity of oncology staff to deliver responsive care. Firstly, assuming that every patient will need the same amount of appointment time risks poor experiences for those left waiting, which is particularly difficult for people with dementia. A rushed approach to appointments means that patients may not be able to take full part in decision-making, as people with dementia may need more time to consider information. Secondly, the single-disease focus is problematic for patients with multimorbidity. Our findings show it is difficult for people to raise issues that are not about cancer, and for staff to respond effectively. Poor outcomes may result because services do not address all the relevant needs and how they interact with each other.

There are potential solutions to reduce tension between the desire for efficiency and the requirement for responsiveness. Our findings suggest that strategies could include identifying in advance people with dementia so that arrangements can be made for them; longer clinic appointments; and geriatric oncology clinics where specialties work together. Practitioners need resources, discretion and autonomy to be able to act outside of the standardised model but are usually constrained. However, as shown by this study, some departments (such as the radiotherapy department who made effective use of dementia champions) were able to act differently, suggesting that change is possible at team level given the right conditions.

### Limitations and strengths

The study was suspended during the first wave of the COVID-19 pandemic. After re-starting, it was challenging to recruit patient and caregiver participants; fewer patients were attending hospital, and the primary researcher (X) was unable to spend time with patients to build rapport. It was not possible to conduct observation, as no non-essential persons could accompany patients during treatment. However, the study protocol was altered to allow for telephone contact and interviews. The relatively low number of formal patient interviews was to some extent mitigated by the inclusion of alternative data collection methods.

## Conclusion

This study contributes to the evidence base around support for people with dementia having cancer treatment. It has offered suggestions for practical and cultural modifications to increase the responsiveness of services, aiming to improve the quality of health services, and decrease health inequalities. Further work is needed to:

- Identify context-specific tools for eliciting the needs of patients and caregivers;
- Embed principles of personalised care in cancer-dementia services;
- Establish and test interventions to target appropriate resources, and enhance autonomy and independence of frontline practitioners.

## Data Availability

All data produced in the present study are available upon reasonable request to the authors.

